# Emergence and Evolution of Big Data Science in HIV Research: Bibliometric Analysis of Federally Sponsored Studies 2000-2019

**DOI:** 10.1101/2021.01.11.21249624

**Authors:** Chen Liang, Shan Qiao, Bankole Olatosi, Tianchu Lyu, Xiaoming Li

## Abstract

**Background:** The rapid growth of inherently complex and heterogeneous data in HIV/AIDS research underscores the importance of Big Data Science. Recently, there have been increasing uptakes of Big Data techniques in basic, clinical, and public health fields of HIV/AIDS research. However, no studies have systematically elaborated on the evolving applications of Big Data in HIV/AIDS research. We sought to explore the emergence and evolution of Big Data Science in HIV/AIDS-related publications that were funded by the US federal agencies.

**Methods:** We identified HIV/AIDS and Big Data related publications that were funded by seven federal agencies from 2000 to 2019 by integrating data from National Institutes of Health (NIH) ExPORTER, MEDLINE, and MeSH. Building on bibliometrics and Natural Language Processing (NLP) methods, we constructed co-occurrence networks using bibliographic metadata (e.g., countries, institutes, MeSH terms, and keywords) of the retrieved publications. We then detected clusters among the networks as well as the temporal dynamics of clusters, followed by expert evaluation and clinical implications.

**Results:** We harnessed nearly 600 thousand publications related to HIV/AIDS, of which 19,528 publications relating to Big Data were included in bibliometric analysis. Results showed that (1) the number of Big Data publications has been increasing since 2000, (2) US institutes have been in close collaborations with China, Canada, and Germany, (3) some institutes (e.g., University of California system, MD Anderson Cancer Center, and Harvard Medical School) are among the most productive institutes and started using Big Data in HIV/AIDS research early, (4) Big Data research was not active in public health disciplines until 2015, (5) research topics such as genomics, HIV comorbidities, population-based studies, Electronic Health Records (EHR), social media, precision medicine, and methodologies such as machine learning, Deep Learning, radiomics, and data mining emerge quickly in recent years.

**Conclusions:** We identified a rapid growth in the cross-disciplinary research of HIV/AIDS and Big Data over the past two decades. Our findings demonstrated patterns and trends of prevailing research topics and Big Data applications in HIV/AIDS research and suggested a number of fast-evolving areas of Big Data Science in HIV/AIDS research including secondary analysis of EHR, machine learning, Deep Learning, predictive analysis, and NLP.

## 1 Introduction

Big Data Science is often referred to as research that capitalizes on the increased volume, variety, velocity, and veracity of data. The proliferation of massive health-related Big Data characterized by high dimensionality and complexity as well as the utilization of advanced statistical and computational technologies offers invaluable opportunities to improve the quality and efficiency of healthcare [1,2]. There has been an increasing consensus in the field that the Big Data science approach will continue to advance our understanding of disease prevention, identification, control, and treatment for decades to come and will be the key to reduce the national and global disease burdens, including those of HIV [3–5].

It is estimated that 37.9 million people are living with HIV (PLWH) worldwide [6]. The HIV treatment cascade, a continuum from prevention, diagnosis, linkage to care, medication adherence and retention to viral suppression (and well-being beyond the viral suppression), is included in the framework for the Joint United Nations Programme on HIV/AIDS (UNAIDS) 90-90-90 targets [7]. The response to the HIV epidemic is complicated by the persistent and prevalent stigma and discrimination against PLWH, socio-cultural barriers to healthcare access, and challenges of disease management (e.g., medicine resistance, comorbidities), all of which necessitate informed decisions and evidence-based strategies for HIV prevention, treatment, and care. Big Data Science allows researchers to innovatively utilize data collected through diverse platforms and within existing systems to understand HIV transmission, clinical outcomes (e.g., disease progression and comorbidities), and physical and psychosocial well-being. The increasing utilization of Big Data can better inform effective policy and practices regarding HIV prevention, treatment, and care.

Big Data Science can be used in exploring HIV transmission and prevention concerning health disparity in HIV infection, and socio-behavioral factors associated with HIV risk. In 2018, there were 37,968 new diagnoses in the US, among which 43% were Black/African American men and 26% were Hispanic/Latino. New diagnoses have increased by 71% in Native Hawaiian/Other Pacific Islanders. The Southern US has 38% of the population yet account for 51% of new diagnoses [8], with a 24% higher proportion of new diagnoses in suburban and rural areas as compared to other regions in the US [9]. Over the last decade, HIV diagnosis rate decreased by 17% in the Northeast and 6% in the Midwest but remains high in the South [10,11]. These findings collectively suggest that factors influencing HIV transmission among vulnerable populations are inherently complex and are involved with nuances from both clinical care and social care. Therefore, a novel approach is demanded to identify the dynamics of HIV transmission through integrating PLWH’s clinical data [e.g., data from Electronic Health Records (EHR)], social care data (e.g., data from community care facilities, mental health, rehabilitation), social determinants of health (SDOH), and health administrative data (e.g., medical claims) [12]. Identifying socio-behavioral factors that are associated with HIV risk appears to be interesting as well. The proliferation of social media, personal health records (PHR), and other forms of patient-generated health data (PGHD) in research has suggested a new role of perceptions and risk behaviors (e.g., stigma, mental status, social support, and SDOH) [13–15].

Big Data Science can also be employed in addressing challenges in HIV treatment and care, such as HIV-associated comorbidities, coinfections, and complications that lead to worse clinical outcomes and high healthcare expenditure [16–21]. However, because relevant studies were often conducted on small and single cohorts with limited variables, the findings were inconclusive [22]. Recently, the detection of HIV-associated comorbidities, coinfections and complications, and evidence-based clinical decision support (CDS) through multi-centered and longitudinal medical records are becoming increasingly available as advances in EHR interoperability unleashed the potential of secondary use of large-scale clinical data. As many researchers have pointed out, Big Data Science is not a “stand-alone tool” but can be incorporated into an existing clinical “toolbox” to advance clinical care and help end the HIV epidemic by transforming data into fundamental knowledge and better health outcomes.

New opportunities in HIV/AIDS research have been created along with the increasingly available sources of Big Data. One major source of Big Data is genomics data that underpin the discovery of genome functions and the genetic basis of AIDS. Since the completion of the Human Genome Project in 2003, next-generation sequencing data have become exponentially available in National Center for Biotechnology Information (NCBI) and have been used in HIV/AIDS research with a focus on HIV mechanism and pathogenesis [23]. Another major source of Big Data is EHR which was incentivized by the Health Information Technology for Economic and Clinical Health Act (HITECH) enacted in 2009 and the Medicare and Medicaid EHR Incentive Programs initiated in 2011 for enhancing the EHR interoperability. A unique role of EHR lies in the capacity of supplying compressive patient data across the continuum of care, which is highly responsive to the continuum of AIDS care [4,24].

However, opportunities and challenges are not apart. Health data exist in a wide variety of formats, with uneven quality and potential bias, which is a critical challenge for HIV/AIDS research because factors contributing to HIV transmission and medical care adherence are from multiple data sources (e.g., clinical, social care, and administrative data). Efforts towards integrated data have been stalled by the lack of comprehensive data repositories [25] and the lack of tailored methodologies in Big Data Science. For example, there have been limited but highly demanded studies in EHR modeling, particularly, modeling temporal clinical events [26,27], data extraction from clinical notes [27], and imputation for missing or sparse data [28].

Over the last two decades, these emerging research topics continued to evolve and jointly called for a combined approach to leveraging Big Data Science for integrating, analyzing, and understanding heterogeneous health data for HIV/AIDS research. Such an effort is consistent with the National Institutes of Health (NIH) initiative of Big Data to Knowledge (BD2K) funded in 2013, in which the notion of “Big Data” was described as an “ever-expanding amount of complex data” in health sciences [1]. Despite an increasing number of publications and funded projects from US federal agencies [25,29,30], no existing studies have systematically reported the implications of Big Data Science in HIV/AIDS research area in terms of research grant support, collaborations, research outcomes, and trending research topics. In this study, we sought to address this literature gap by employing the bibliometric method to analyze publications sponsored by research grants from US federal agencies that focus on Big Data in HIV/AIDS research. The bibliographic analysis is rooted in information science and has been used for detecting networks, patterns, and trends among intricate and large-scale bibliographic metadata [e.g., titles, abstracts, authors, institutes, keywords, Medical Subject Headings (MeSH) terms] [31]. This study focuses on academic research funded by the NIH and other federal agencies and therefore does not include “Big Data” projects done by state and city health departments through other funding sources such as Centers for Disease Control and Prevention (CDC) grants (e.g., data-to-care initiatives) [32]. Our early data mining work has systematically analyzed federal funding allocation on HIV/AIDS research in the US [11]. The present study provides additional insights to extend our knowledge about how federal efforts and research productivity are shaped by Big Data Science. Findings of the study may retrospectively reveal interesting research patterns and predictively inform trending research foci and future research directions in Big-Data-driven HIV/AIDS research.

## 2 Materials and methods

### 2.1 Data extraction

#### 2.1.1 Federally funded HIV/AIDS projects

We identified HIV/AIDS-related projects in NIH ExPORTER. The database consists of administrative data for research projects funded by seven federal agencies: NIH, the Administration for Children and Families (ACF), the Agency for Healthcare Research and Quality (AHRQ), CDC, Health Resources and Services Administration (HRSA), U.S. Food and Drug Administration (FDA), and Veterans Affairs (VA).

Data query with ExPORTER restricts the time of projects to be from 2000 to 2019. Because data before 2020 are stored in CRISP (1970-2009), the legacy system of ExPORTER, with different variables and formats, we did not include data from before 2020. To identify HIV/AIDS-related projects, we used regular expression (“HIV” OR “AIDS” OR “HIV/AIDS” OR “HIV-1” OR “human immunodeficiency virus” OR “acquired immunodeficiency syndrome”) to be applied in the fields of “abstract_text”, “project_terms”, “project_title”, and “study_section_name”. These four fields were selected from a total of 46 fields in ExPORTER. The regular expression was determined by an HIV/AIDS expert (SQ) and a medical informatics expert (CL). The “project_IDs” was used as a unique identifier. Fields “project_terms” and “study_section_name” contain controlled text that was determined by the NIH; “abstract_text” and “project_title” contain free text that was provided by the recipients of funded projects. Therefore, we performed lemmatization and punctuation removal on free-text fields prior to the regular expression. The data processing and extraction were performed using in-house Python programming codes.

#### 2.1.2 Publications involved with Big Data

We extracted publications that were resulted from these funded HIV/AIDS projects by querying about “PMIDs” that were linked to the “project_IDs” we have identified. With these “PMIDs”, we batch-downloaded MEDLINE metadata by interacting with Entrez Programming Utilities (E-utilities) at the NCBI.

To query Big-Data-related PMIDs, two domain experts (CL and SQ) selected a collection of MeSH terms that represent themes of Big Data. See Table 1. These MeSH terms were used in Perl queries for E-utilities. The pseudo-query is as follows:

**Table 1.** Big-Data-related MeSH terms.

| MeSH terms | MeSH IDs |
| --- | --- |
| big data | D000077558 |
| data mining | D057225 |
| data science | D000077488 |
| machine learning | D000069550 |
| deep learning | D000077321 |
| natural language processing | D009323 |
| information storage and retrieval | D016247 |
| data warehousing | D000073458 |
| knowledge discovery | D063369 |
| artificial intelligence | D001185 |
| algorithms | D000465 |
| cloud computing | D000067917 |

*[HIV/AIDS-related PMIDs] AND (“big data” OR “data mining” OR “data science” OR “machine learning” OR “deep learning” OR “natural language processing” OR “information storage and retrieval” OR “data warehousing” OR “knowledge discovery” OR “artificial intelligence” OR “algorithms” OR “cloud computing”)*

The query was applied on the [TW] (text words), a field in MEDLINE metadata that includes free text in the title, abstract, other abstract, MeSH terms, MeSH Subheadings, Publication Types, Substance Names, Personal Name as Subject, Corporate Author, Secondary Source, Comment/Correction Notes, and Other Terms ([OT] field) typically non-MeSH subject terms (e.g., keywords).

Upon the completion of the query, we received bibliographic information of 19,528 publications in MEDLINE format. This total excludes less than 5% of data loss that is due to erroneously reported or downloaded PMIDs [33]. Figure 1 shows the diagram of data extraction procedures.

**Figure 1.**
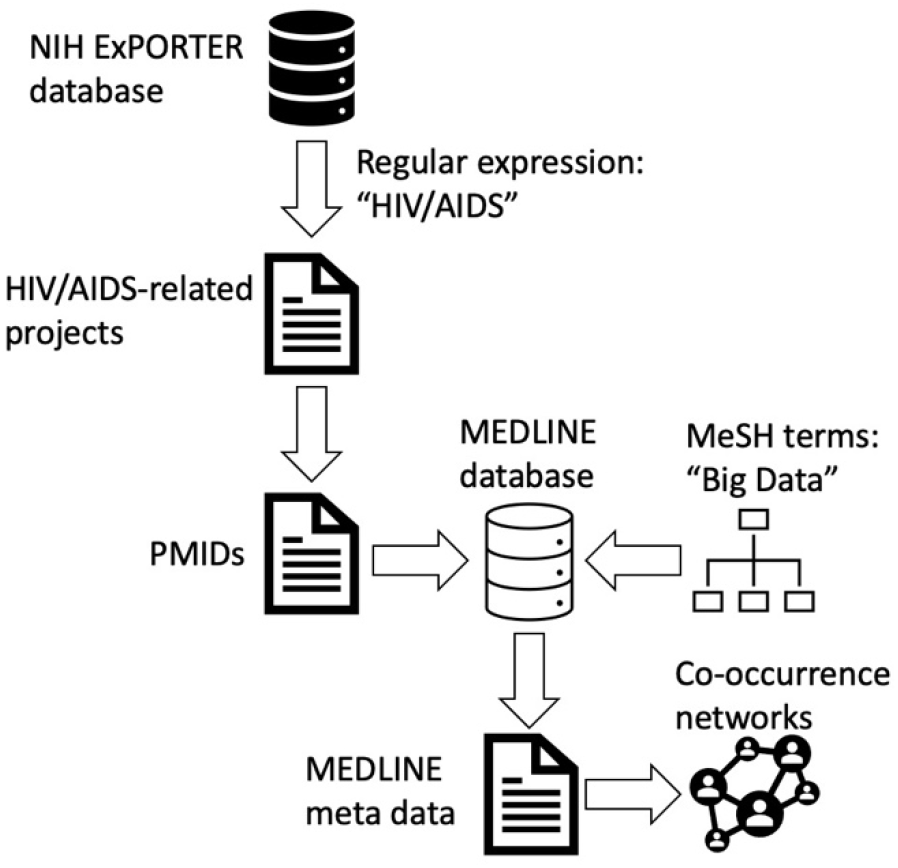
Data extraction flowchart.

### 2.2 Data analysis

We constructed co-occurrence networks of the bibliographic data using the following data elements available in MEDLINE metadata: countries, institutes, MeSH terms, and keywords. These co-occurrence networks exhibit frequency occurrences of the involved data elements over time (2000-2019). For example, we can explore international collaborations by assessing the publications associated with the same countries. Aside from the co-occurrence networks, we also generated clusters for MeSH terms and keywords, which were used to detect salient research topics and trends over time. Specifically, we employed the Latent Semantic Analysis (LSA) [34] and log-likelihood ratio (LLR) for analyzing the neighboring MeSH terms and keywords to be included in a cluster. These methods measure similarities among MeSH terms and keywords. Co-occurrence networks, clustering, and visualization were completed on CiteSpace [35].

## 3 Results

The number of relevant publications has constantly increased from 818 as of 2000 to 2,450 as of 2016 (Figure 2). There was a decline in 2017 (n=2,431), 2018 (n=2,176), and 2019 (n=1,586).

**Figure 2.**
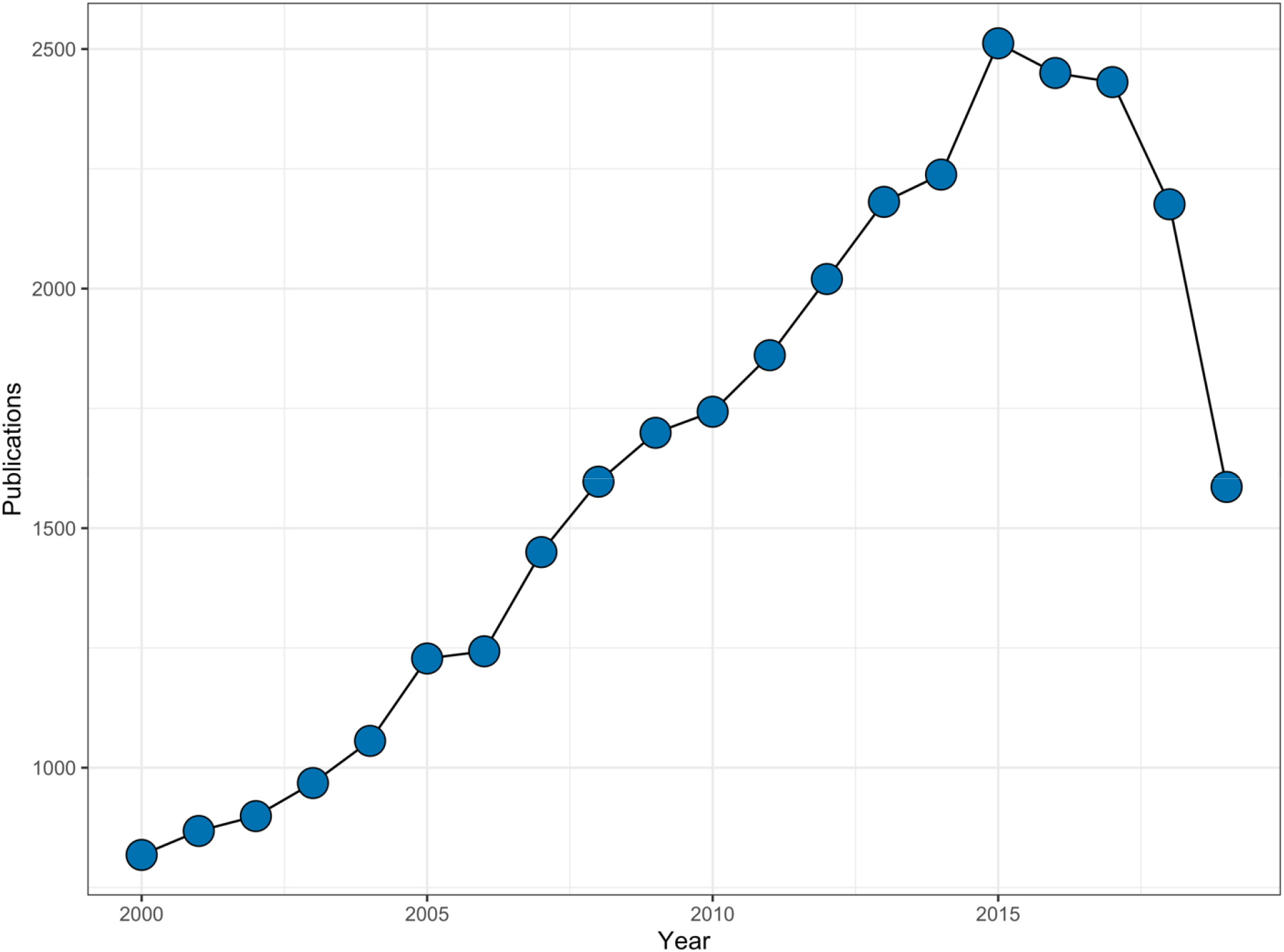
Number of publications by year.

### 3.1 International collaborations

97% (n=18,937) of the publications were published with at least one US institute. US institutes often collaborate with China (n=462, 2.4%), Canada (n=412, 2.1%), and Germany (n=326, 1.7%). See Table 2. The US began to apply Big Data Science in HIV/AIDS research as early as 2000. China, Canada, and France initiated Big Data in HIV/AIDS research earlier than other non-US countries.

**Table 2.** Number of publications by countries.

| Countries | No. of publications | Percentage |
| --- | --- | --- |
| US | 18,937 | 97.0% |
| China | 462 | 2.4% |
| Canada | 412 | 2.1% |
| Germany | 326 | 1.7% |
| France | 256 | 1.3% |
| Australia | 190 | 1.0% |
| UK | 177 | 0.9% |
| Switzerland | 148 | 0.8% |
| Spain | 142 | 0.7% |
| Italy | 133 | 0.7% |
| Total | 19,528 | 100% |

### 3.2 Participation and productivity of institutes

As shown in Table 3, the University of California system (n=309) and Harvard University (n≥284) were among the most productive institutes. Note that research centers, schools, and research institutes affiliated with a university could be counted apart from their parent institutes in MEDLINE data format because authors do not constantly include their parent institutes in publications.

**Table 3.** Number of publications by institutes.

| Institutes | No. of publications |
| --- | --- |
| University of California | 309 |
| MD Anderson Cancer Center | 138 |
| *Harvard Medical School | 134 |
| *Athinoula A. Martinos Center for Biomedical Imaging | 103 |
| University of Pennsylvania | 77 |
| University at Albany | 53 |
| Jude Children's Research Hospital | 53 |
| *Harvard T.H. Chan School of Public Health | 47 |
| Stanford Center for Biomedical Informatics Research | 45 |
| Emory University | 44 |
| Fred Hutchinson Cancer Research Center | 43 |
| Duke University Medical Center | 30 |
| Total | 19,528 |
| * Same parent institute |  |

We developed a time zone figure to visualize the participation and productivity of institutes over time (Figure 3). This figure shows institutes that were not only productive in Big-Data-related HIV/AIDS research but also entered the field early, such as the University of California system, MD Anderson Cancer Center, and Harvard Medical School. Several institutes were not at an advantage in terms of the number of publications but started exploring the relevant research as early as between 2000 to 2008, including Johns Hopkins University School of Medicine, Drew University, Fred Hutchinson Cancer Center, Jude Children’s Research Hospital, Massachusetts General Hospital, etc. Despite a fairly large number of HIV/AIDS research coming from the field of public health, the only public health institute that was active in Big-Data-related research was Harvard T.H. Chan School of Public Health as dated in 2015 (Figure 3), suggesting that in public health Big Data was introduced to HIV/AIDS research comparatively late.

**Figure 3.**
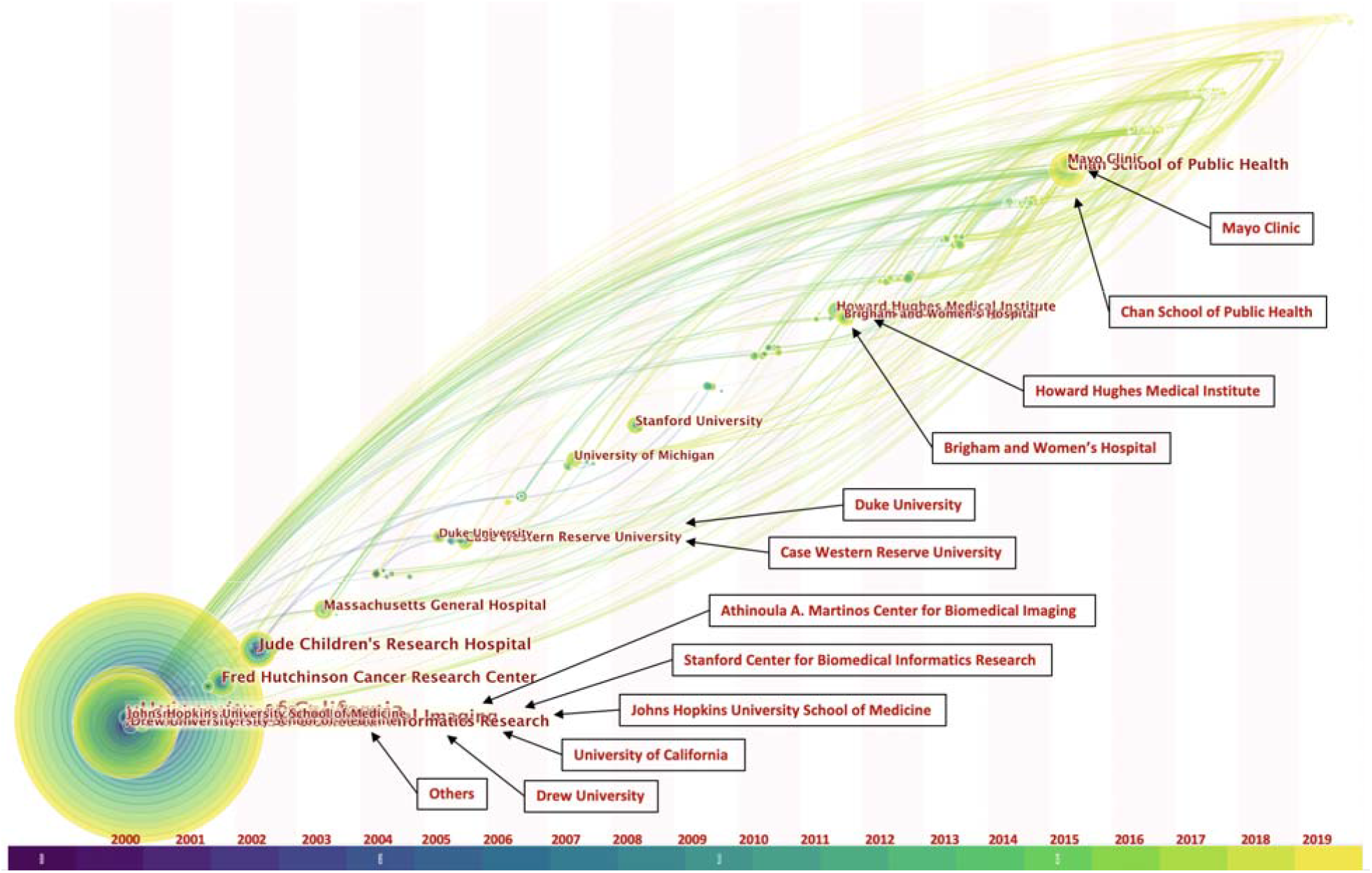
Time zone figure. Each node shows the inception year of an institute measured by the number of related publications. The size of nodes indicates the frequency of relevant publications by institutes. The colors indicate the years by which an institute started to produce significant amounts of relevant publications, spanning 2000 (dark) to 2019 (light). The edges indicate the collaborations between institutes. For clarity of the figure, the thickness of the edges is unified and does not represent frequency differences. The time zone where an institute displays indicates the year the institute began to produce relevant publications.

### 3.3 Areas of study

Figure 4 demonstrated the top 100 bursts of MeSH terms and keywords, in which bursts measure the period of time that related MeSH terms and keywords rise sharply in frequency in the co-occurrence networks [36]. These bursts carried critical implications in terms of emerging research topics and methods in publications. In addition to MeSH terms and keywords that have been used in high-frequency throughout 2000-2019, we found several MeSH terms and keywords that did not emerge until recent years. Some mentions of medical conditions are not directly associated with HIV/AIDS because they are comorbidities and complications discussed in the same papers that HIV/AIDS topics are included. To ease the description, we organized them into the following categories. (1) Machine learning methods: “deep learning”, “convolutional neural network”, “feature selection”, “random forest”, “clustering”, “LASSO”, “sensitivity”, “specificity”, “Markov Chain Monte Carlo”, “cross validation”, “support vector machine”, etc. Arguably, these methods and techniques have been increasingly used for automated classification and clustering of health data. (2) Emerging health data related terms: “data sharing”, “electronic medical record”, “missing data”, “imputation”, “social media”, etc. These terms provide insights into the growing number of electronic health data and multi-modal health data that have been used for Big Data analysis. (3) Biomedical techniques and related topics: “precision medicine”, “RNA-seq” (RNA sequencing), “radiomics”, “parallel imaging”, “diffusion MRI”, “fMRI”, “genomics”, “tractography”, “network analysis”, “imaging analysis”, “next-generation sequencing”, “compressed sensing”, “flow cytometry”, “signal processing”, “imaging processing”, “mass spectrometry”, etc. (4) HIV-associated comorbidities, coinfections, and complications: “glaucoma”, “cancer”, etc.

**Figure 4.**
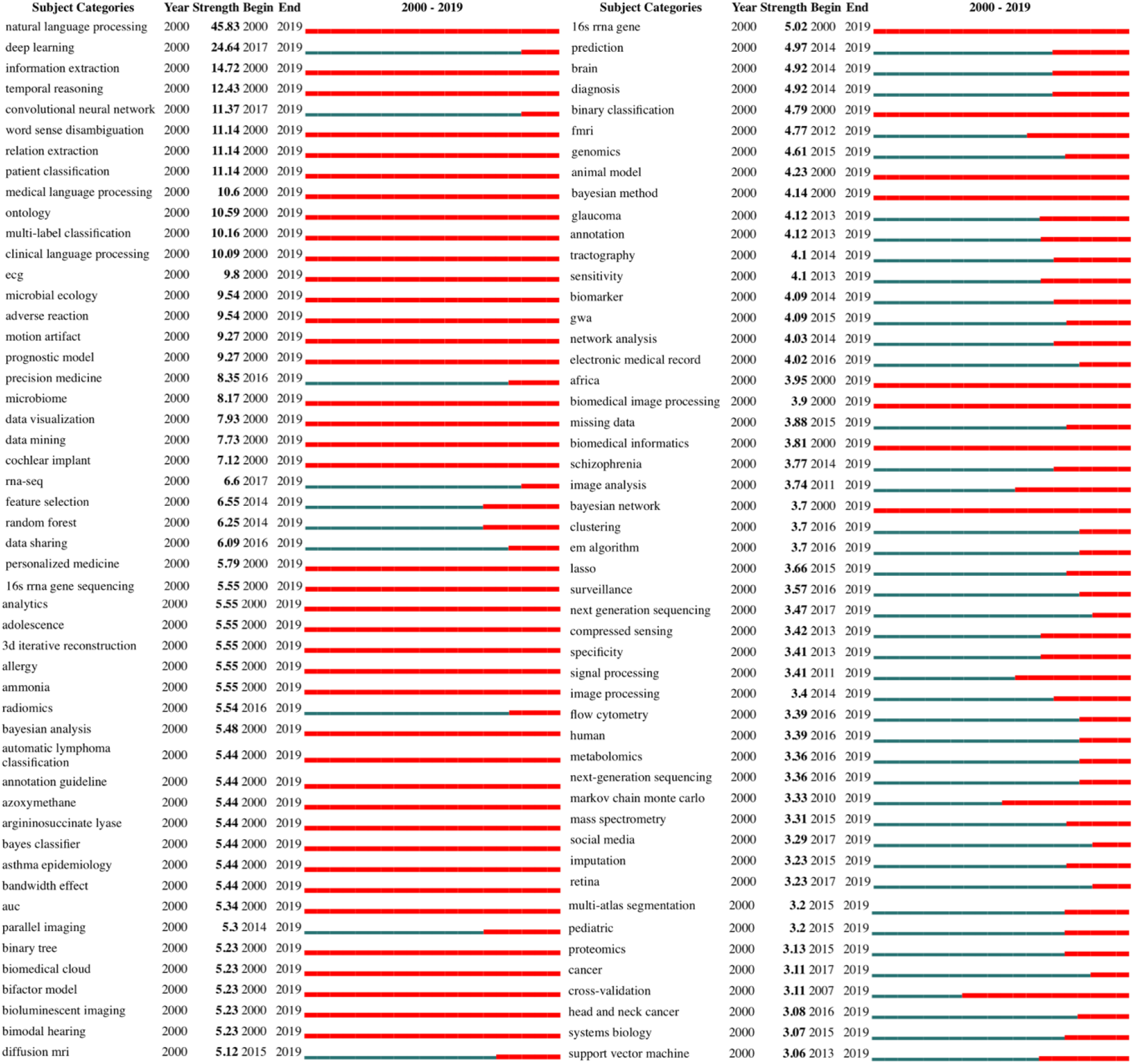
Top 100 MeSH term and keyword bursts. The red bar indicates the course of bursts over time.

On top of the co-occurrence network of MeSH terms and keywords, we generated clusters of MeSH terms and keywords. Table 4 shows the valid clusters, surrounding terms included in the clusters, and cluster parameters. The validity of the clusters was measured by Silhouette scores, in which a high score indicates a high consistency of terms within a cluster [37]. These clusters were generated by LLR. The LAS algorithm failed to produce robust results with low consistency in the clusters. Figure 5 visualizes these colored-labeled clusters with colored nodes of MeSH terms and keywords. The labeled names of clusters in Figure 5 are the terms with top log-likelihood ratio in a cluster and do not necessarily represent the meaning of a cluster. The meaning of a cluster is interpreted by domain experts (CL, SQ) in the review of the included terms. To the best of our understanding, cluster #7 is related to eye disorder, comorbidity with AIDS. Cluster #6 is related to hearing loss, another comorbidity with AIDS. Cluster #5 is related to clinical natural language processing (NLP) which is focused on integrating and mining clinical notes within the electronic health records (EHR). Cluster #4 is related to the methods of structural biology. Cluster #3 is related to breast cancer, one of the comorbid cancers with AIDS. Cluster #2 is related to genomics, a field of biology focusing on genomes. Cluster #1 is comparatively less coherent as it involves medical imaging, clinical NLP, and comorbidity. Cluster #0 is related to diagnostics and clinical care of AIDS. HIV-associated comorbidities generally appear across most of the clusters.

**Table 4.**
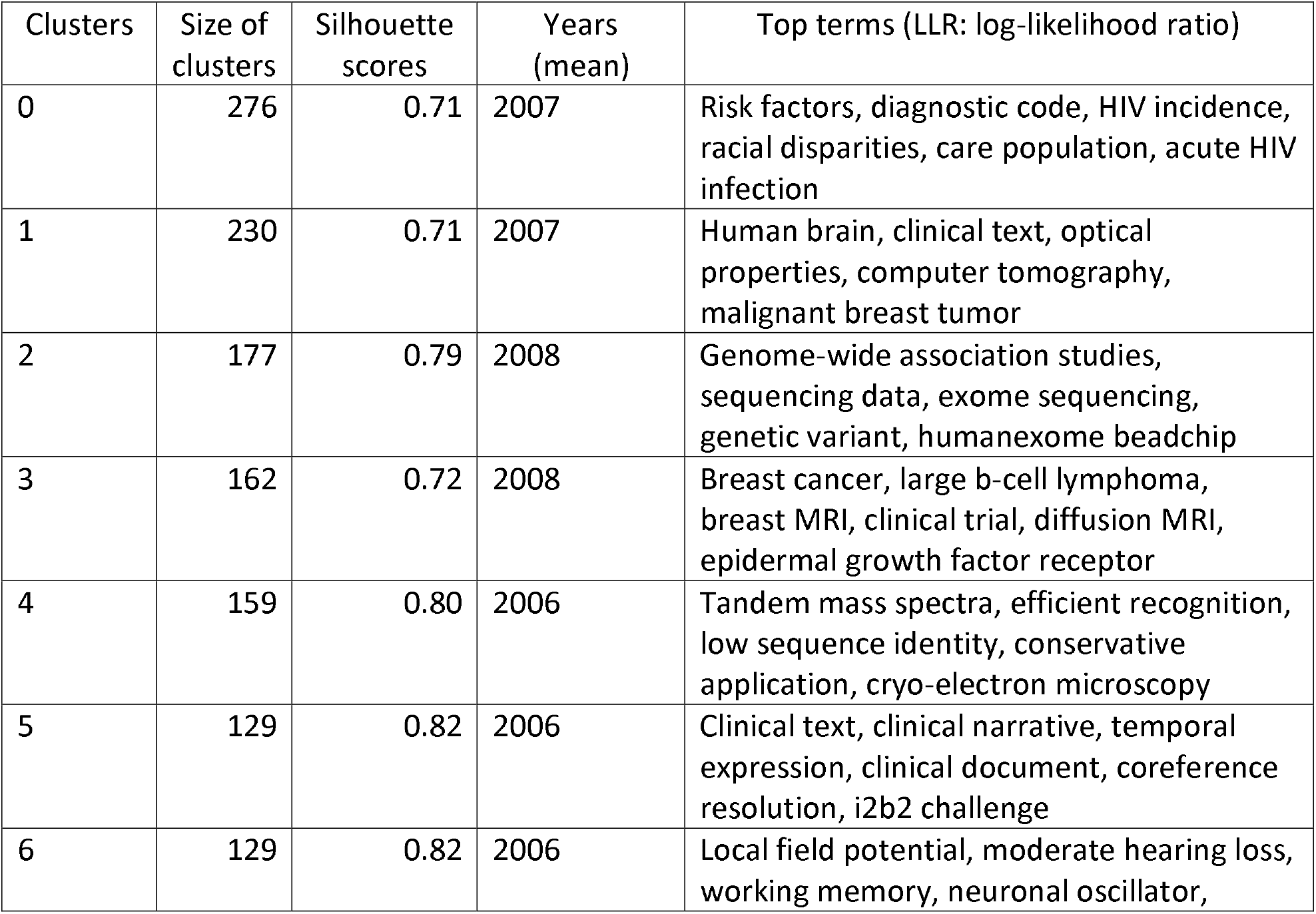
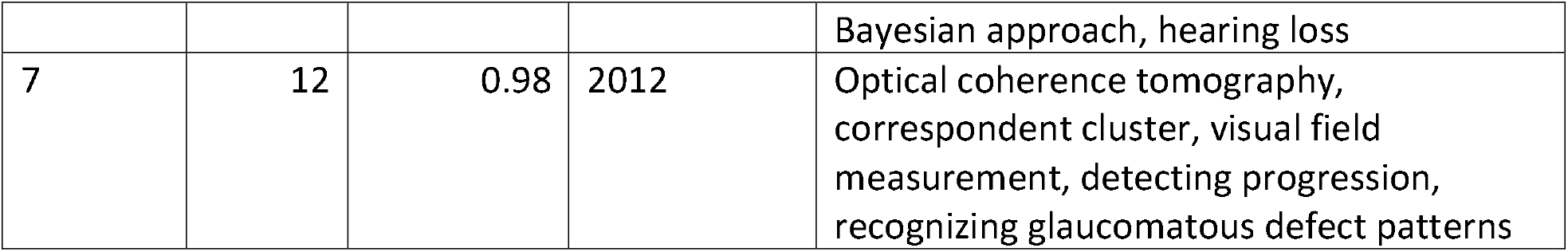
Clusters generated based on the co-occurrence network of MeSH terms and keywords.

**Figure 5.**
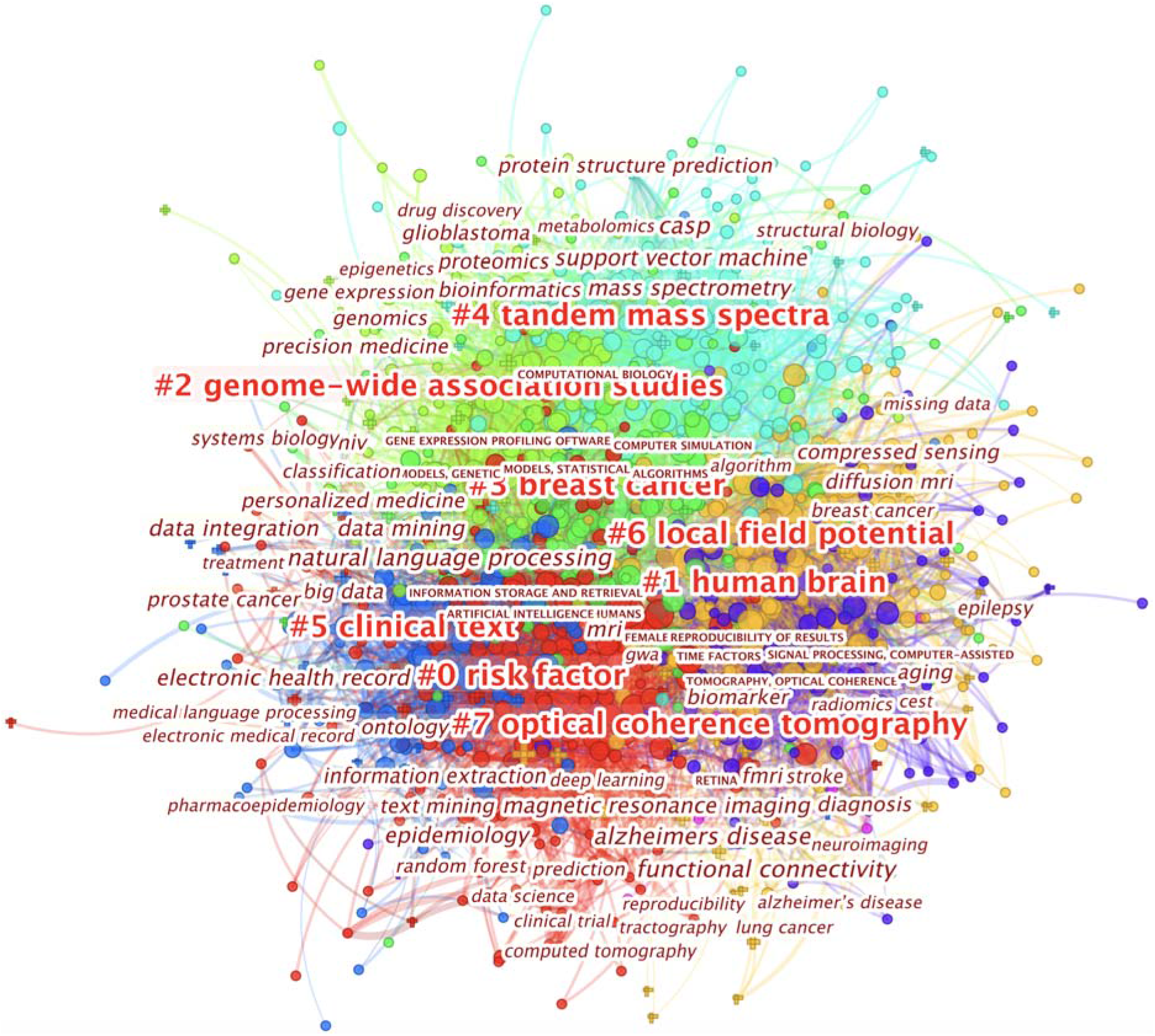
Visualization of clusters. Clusters are color labeled and annotated by a name (red) that refers to the term of top log-likelihood-ratio in a cluster. Other terms within the clusters are in maroon. The clusters are ranked by the Silhouette scores, in which a high value indicates well-coherent terms to its own cluster.

To visualize the temporal dynamics of the detected clusters and the surrounding terms, we projected the clusters onto a chart with a timeline view (see Supplement). This visualization demonstrated a number of findings that were not revealed in other analyses, e.g., terms within a cluster appearing at different times. Notably, machine learning algorithms, Deep Learning, and CDS raised rapidly in conjunction with the emergence of EHR-related and patient-data-related topics. Many of such combinations were related to HIV-associated comorbidities, coinfections, and complications. Among the eight clusters, at least three (clusters #7, #6, and #3) were directly related to HIV comorbidities, and cluster #7 was the only one that disappeared since 2017.

## 4 Discussion

We extracted Big Data publications based on HIV/AIDS research grants that were funded by the US federal agencies from 2000 to 2019. We further demonstrated the emergence and evolution of diverse research topics, methods, data sources, and research collaborations in these publications. Our study captured a consistent upsurge in the number of both US federal funding support and the resulted publications for Big-Data-related HIV/AIDS research over time, suggesting an increasingly important role of Big Data Science in HIV/AIDS research. The exception is the decline from 2017 to 2019 that is likely related to the budget cut on federal funding in HIV/AIDS research and is related to the incomplete integration of new data in ExPORTER as found in other studies [38]. This trend is consistent with the US federal funding for HIV/AIDS research measured by total cost in the same period of time [11,39].

Analysis of international collaborations showed that these US-funded projects have fostered close and productive collaborations with countries such as China, Canada, Germany, and France, measured by the number of co-authored publications. These international collaborations started as early as 2000. With regard to institutional productivity, the University of California system, MD Anderson Cancer Center, Harvard University, University of Pennsylvania were among the most productive institutes. We have three major findings regarding the temporal dynamics. First, institutes (e.g., the University of California system, MD Anderson Cancer Center, Harvard Medical School) that applied Big Data in HIV/AIDS research as early as 2000 have been highly productive over the last two decades and have established wide collaborations with other institutes early. Some institutes (e.g., Johns Hopkins University School of Medicine, Drew University, Fred Hutchinson Cancer Center, Jude Children’s Research Hospital, Massachusetts General Hospital) that initiated Big-Data-related HIV/AIDS research as early as around 2000 were comparatively less productive probably because of the diverse research strategies and relatively small setting as compared to large academic settings such as the University of California system. Second, most of the high-profile institutes were centered on biomedical and clinical research. Third, public health communities have been making substantial contributions to HIV/AIDS research over time but were not active in Big Data until a much later time marked by the emergence of Harvard T.H. Chan School of Public Health in 2015.

The emergence of Big Data Science in HIV/AIDS research was as early as 2000 but the research foci and context have changed substantially over time. The application of Big Data Science was found in the following cross-cutting themes: (1) HIV-associated comorbidities, coinfections, and complications. Big Data methods adopted in this theme varied by specific research topics. For example, Markov Chain and Stochastic processes were widely used in modeling cancer progression (e.g., breast cancer) as early as 2001. Survival analysis, genetic testing, and bioinformatics were introduced a few years later. In recent years, an increasing number of studies within the theme were found in conjunction with the secondary use of EHR. (2) System biology including genomics and bioinformatics. This theme focused on fundamental research questions such as HIV/AIDS mechanism, pathogenesis, and antiretroviral therapy (ART) drug towards HIV cure and next-generation therapies. (3) Secondary use of EHR. This theme widely interacted with HIV-associated comorbidities, coinfections, and complications in terms of modeling clinical characteristics based on large-scale EHR. The theme interacts with methods such as machine learning, Deep Learning, predictive analysis, and NLP. It also interacted with epidemiological, behavioral, and social aspects of HIV/AIDS research. (4) Increased utilization of PGHD and social media data in research. Although this theme is relatively small in the number of publications, it grew rapidly in recent years. Additionally, there are other cross-cutting topics including pharmacy, therapies, medical imaging, etc. The emergence of these research themes is generally aligned with (and responsive to) the evolving trend and priorities of HIV prevention and treatment cascade as well as the development of analytic techniques and the increased availability of data sources [4,12,24,40].

This study is subject to limitations. First, the search strategy for identifying relevant projects from ExPORTER favors minimizing miss-detected relevant projects and, as a result, may tolerate a small number of projects that include but do not primarily focus on HIV/AIDS. Second, our bibliographic data did not include citations because the data were from MEDLINE. Third, our method is limited in capturing meaningful clusters that have a small number of publications because bibliographic analyses heavily rely on co-occurrence probabilities. Fourth, detecting a full spectrum of Big Data related methods and datasets is important but requires comprehensive NLP analyses, which is beyond the scope of this study. Fifth, because bibliometric analysis processes the entire corpus of publications, it may not be intuitive for audiences with respect to sample size, inclusion/exclusion criteria of publication selection, and other typical information a traditional systematic review study would be able to provide in a standard CONSORT flow diagram. Sixth, because our findings are based on federally funded academic research, they do not reflect service-based research funded by agencies such as CDC.

## 5 Conclusion

This study is the first that systematically analyzed the emergence and evolution of applying Big Data Science in HIV/AIDS research, which provided critical implications on US federal research priorities, trending research topics, and challenges. This systematic bibliometric analysis contributes to the discussions of future study direction and informs effective strategies for interdisciplinary and cross-institutional collaborations so that the implications of Big Data Science can be better integrated into the efforts to end the HIV epidemic.

## Data Availability

Data referred to in the manuscript are available upon request.

## Acknowledgement

Research reported in this publication was supported by the National Institute of Allergy and Infectious Diseases of the National Institutes of Health under award number R01AI127203.

## Authors’ contributions

Conception and design of study: CL, SQ, XL.

Acquisition of data: TL, CL, SQ.

Analysis and/or interpretation of data: CL, SQ, XL.

Drafting the manuscript: CL.

Revising the manuscript critically for important intellectual content: XL, SQ, BO.

Approval of the version of the manuscript to be published: all authors.

## Summary table

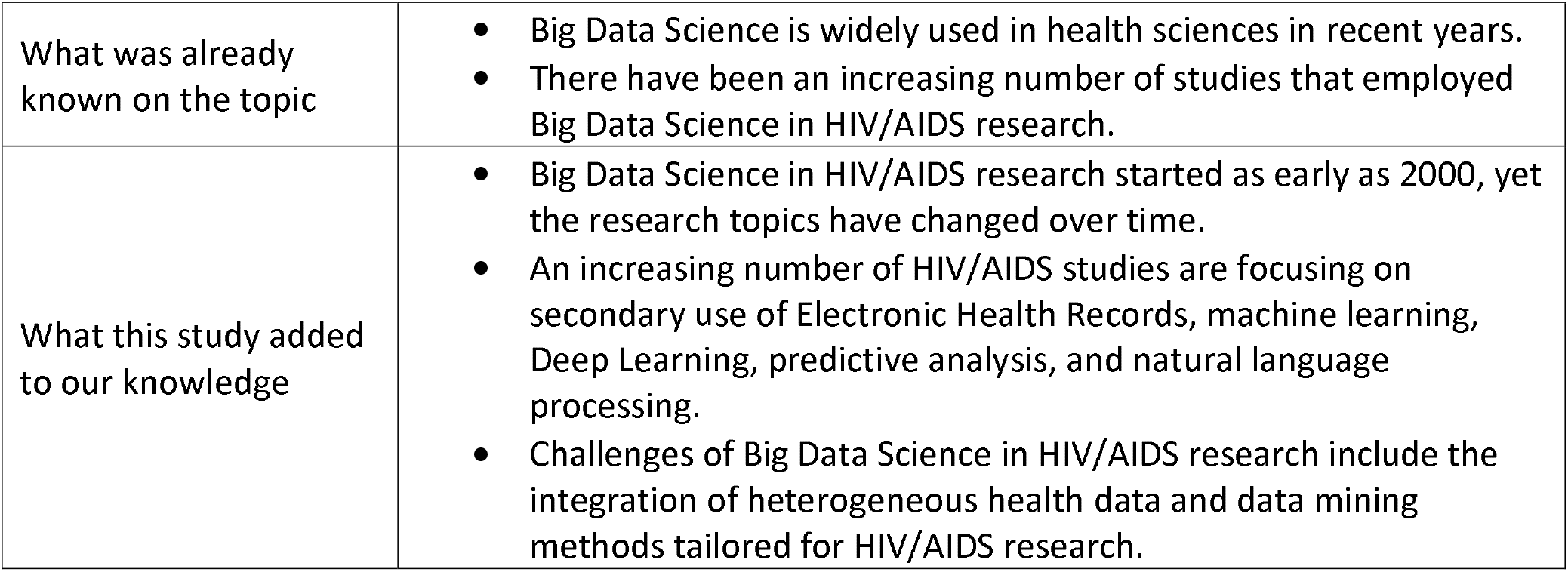

---

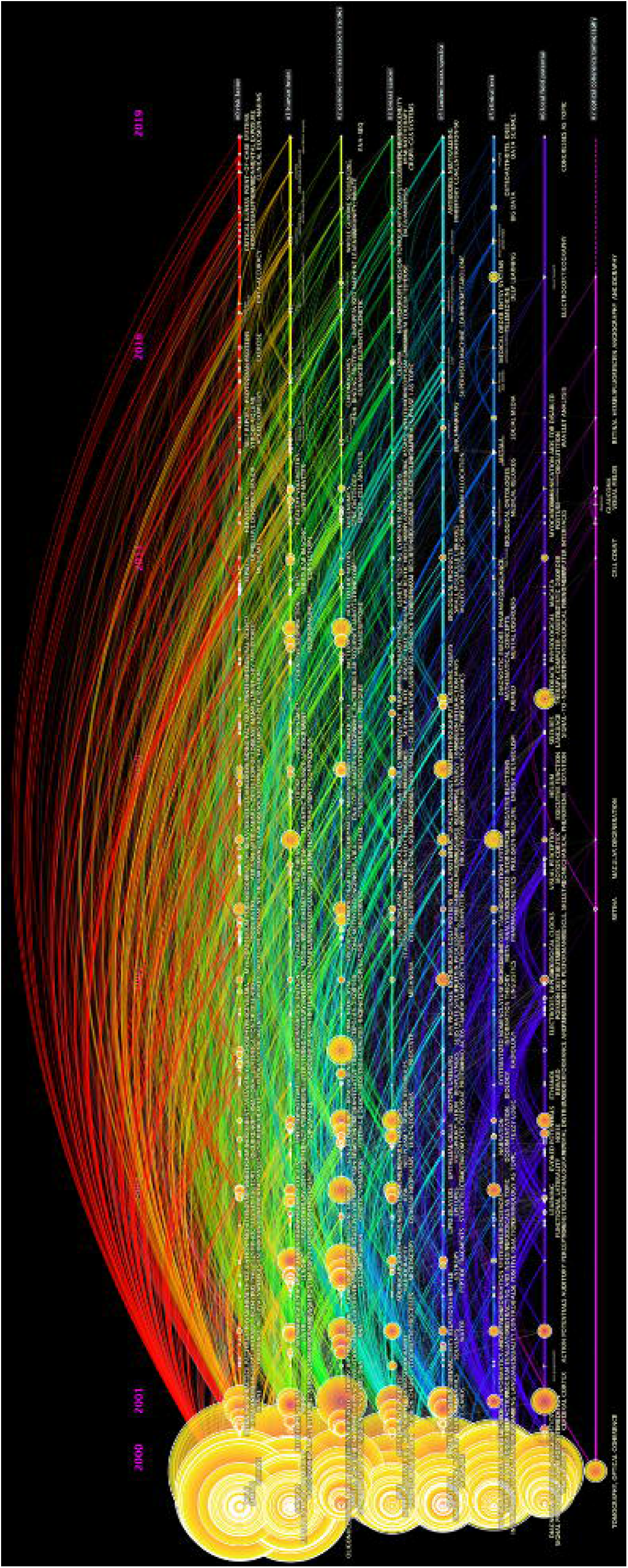

